# Empirical Model of Spring 2020 Decrease in Daily Confirmed COVID-19 Cases in King County, Washington

**DOI:** 10.1101/2020.05.11.20098798

**Authors:** Jared C. Roach

**Affiliations:** Institute for Systems Biology, Seattle, WA

**Keywords:** R_0_, reproduction number, reproduction ratio, reproduction rate, reproductive number, reproductive ratio, reproductive rate, COVID-19, SARS-CoV-2, half-life, SIR, SEIR, SEIRS, compartmental model, outbreak, epidemic, pandemic

## Abstract

Projections of the near future of daily case incidence of COVID-19 are valuable for informing public policy. Near-future estimates are also useful for outbreaks of other diseases. Short-term predictions are unlikely to be affected by changes in herd immunity. In the absence of major net changes in factors that affect reproduction number (R), the two-parameter exponential model should be a standard model – indeed, it has been standard for epidemiological analysis of pandemics for a century but in recent decades has lost popularity to more complex compartmental models. Exponential models should be routinely included in reports describing epidemiological models as a reference, or null hypothesis. Exponential models should be fitted separately for each epidemiologically distinct jurisdiction. They should also be fitted separately to time intervals that differ by any major changes in factors that affect R. Using an exponential model, incidence-count half-life (*t*_1/2_) is a better statistic than R. Here an example of the exponential model is applied to King County, Washington during Spring 2020. During the pandemic, the parameters and predictions of this model have remained stable for intervals of one to four months, and the accuracy of model predictions has outperformed models with more parameters. The COVID pandemic can be modeled as a series of exponential curves, each spanning an interval ranging from one to four months. The length of these intervals is hard to predict, other than to extrapolate that future intervals will last about as long as past intervals.

## INTRODUCTION

Projections of the near future of daily case incidence during epidemics are valuable for informing both individuals and public policy. Simple epidemiologic theory predicts an exponential change in viral incidence in a largely susceptible population, in which relatively few individuals are immune (Ross, 1915). In 1928, Merritte Ireland reported for the 1918 influenza pandemic, “If this was really the beginning of the great epidemic wave one should expect that if these series of data were plotted out on a logarithmic scale the increase from week to week would plot out as a straight line following the usual logarithmic rise of an epidemic curve” (Ireland, 1928). The basic insight that outbreaks of infectious disease should fit an exponential model remains valid a century later. An exponential model is simultaneously simple and mechanistic. There are a mere two parameters, yet these parameters enable modeling a biologically and epidemiologically plausible mechanism. In short, a fixed rate of transmission between individuals results in an exponential change in case counts. Furthermore, the parameters of the model are interpretable. In particular, the incidence-count half-life (*t*_1/2_) has a specific, useful, and understandable meaning. Together, these five factors – historical acceptance, simplicity, plausibility, interpretability, and utility – make the exponential model best suited as the default model by which all other models should be compared.

## RESULTS

A model for King County during Spring 2020 is presented here as an example of exponential model fitting. The fit interval is bounded by a start date representing widespread adoption of physical distancing public policy on March 26, 2020, and an end date of June 17, 2020 (near the end of the first wave). It spans a time interval of 82 days, or several months. The March 26 bound was selected for two reasons. First, the reproduction number (R) of SARS-CoV-2 will have changed dramatically after physical distancing (aka, “social distancing”) policies were implemented in King County earlier in March. For example, Public Health — Seattle & King County (PHSKC) began encouraging physical distancing on March 10 and Washington State issued a “stay at home” directive on March 23. The inflection point of daily case counts of March 26 chosen as a bound was not caused by a rapid change in herd immunity. It was caused by a dramatic change in R secondary to intentional changes in human behavior. This cause is markedly different in character than the cause of the inflection point in most classical models (e.g., SIR), which is due to saturation of susceptible individuals. A May 2020 estimate of cumulative infections and therefore likely fraction of immune individuals in King County is only 2.1% (Thakkar, Burstein, Klein, and Famulare, 2020), and this number has been slow to change. Therefore, an unmodified Susceptible, Infected and Recovered (SIR) model is not appropriate for this situation. Second, inspection of the raw data suggests an inflection point at or around this date. The highest confirmed case count to date occurred on April 1, reported as 219 case counts, but there is sufficient variability in the daily reported case counts that one cannot rule out a true peak of the curve occurring either several days earlier or later. The analysis is robust to slightly different choices of threshold for date, including any date from March 25 to April 3, representing the peak of the Spring 2020 COVID-19 outbreak in King County (also known as “the first wave”). Visual inspection of the raw data after March 26 also suggests that an exponential fit would be excellent.

Nonlinear regression was performed on the daily confirmed case counts (**Figure 1**), with excellent fit (R^2^ = 0.87). The equation for the fitted curve is, where *N* = predicted daily confirmed cases and *t* = days after March 26:

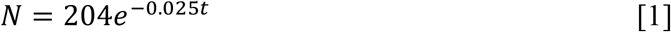

**Figure 1.**
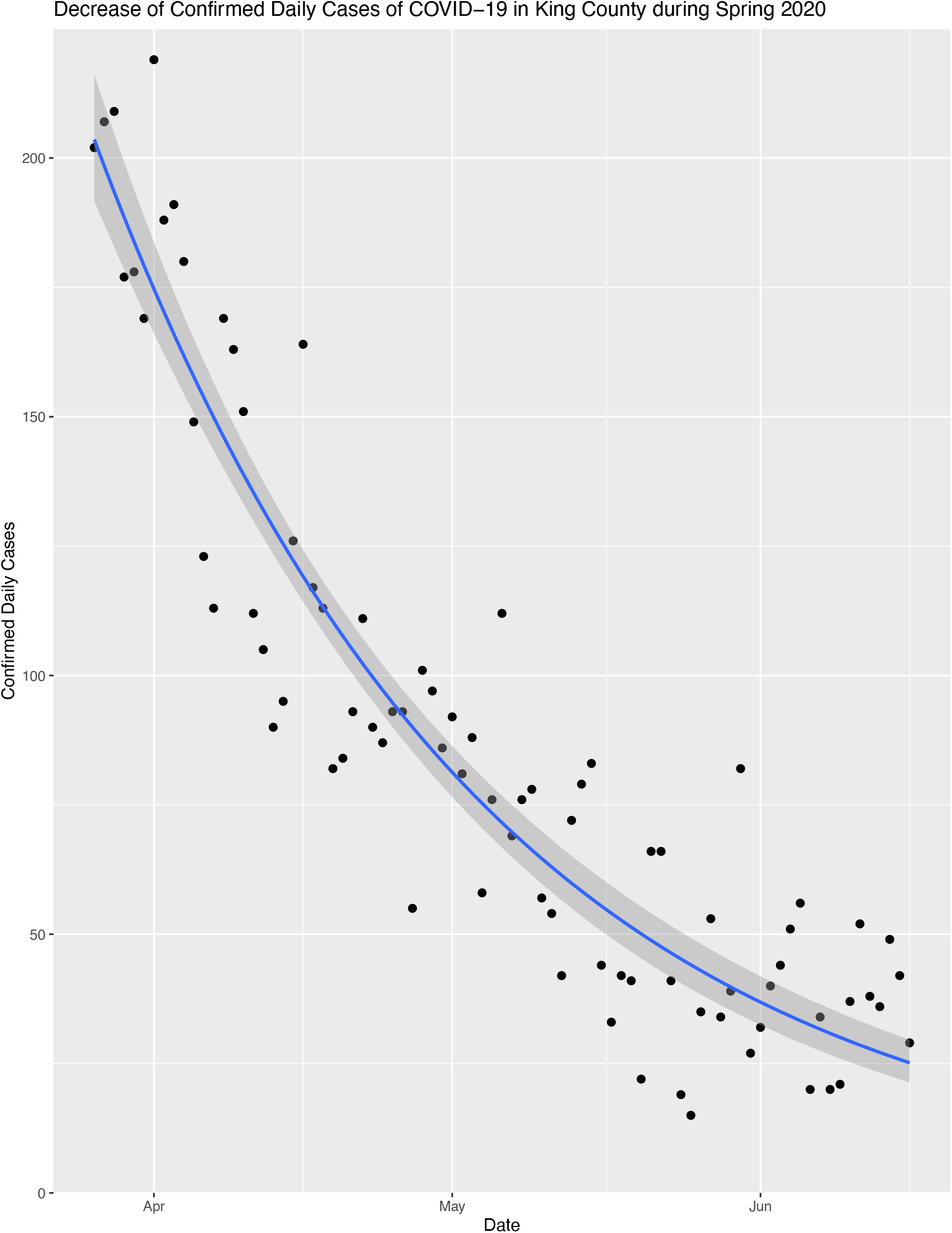
Exponential Fit to Model Decrease of Confirmed Daily Cases of COVID-19 in King County during Spring 2020. Each point is the number of confirmed cases of COVID-19 for each day as reported on June 17, 2020 by PHSKC. The date range is March 26 to June 16, 2020.

The two parameters of the model are the initial case count (*N*0 = 204) and a rate constant (*λ* = 0.025). Consequently, the half-life (*t*_1/2_) is

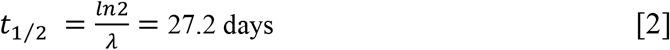

If the case count were increasing, the half-life would be instead described as a doubling time. The King County model fit has a half-life (*t*_1/2_) of 27.2 days (95% CI: 24.6—30.1). That is, the number of daily confirmed cases is expected to drop by 50% every 27 days. With approximately 80 cases observed on May 2, it predicted about 40 cases per day on May 29, 20 cases per day on June 25, and 10 cases per day on July 22, 2020. The Institute for Disease Modeling (IDM) model for COVID-19 assumes 4-day latency after infection followed by a period of 8-days of uniform infectiousness (Thakkar, Burstein, Klein, Schripsema, and Famulare, 2020b). With these parameters, the model predicts R = 0.82 (95% CI: 0.80—0.83). The combination of simple model, large number of data points, and low residuals lead to narrow confidence intervals for these analyses.

One test of a model’s utility is its ability to predict the future. Predictions from our model were first made on May 3 and ensconced in MedRxiv on May 11, 2020. The model parameters have remained stable and the predictions have largely been borne out. However, sub-analyses suggest that R has been increasing since March 26, slowly at first and more rapidly in weeks closer to the end of May (**Table 1**). Alternatively, or in conjunction, increases in testing have enabled a higher ratio of confirmed to true case counts. It is difficult to establish from these data alone that these changes in R are significant (see **Figure S1**); they could result from a combination of progressive improvement in PHSKC’s estimates of previous days’ case counts, increased number of training data points, and noise. Indeed, a previous version of this paper included a correction for testing rate that at least partially accounts for this apparent increase in R (Roach, 2020). However, an increase in R has been expected due relaxation of public health policies and increases in mass gatherings, so it is likely that these observations reflect a true increase in R.

**Table 1.**
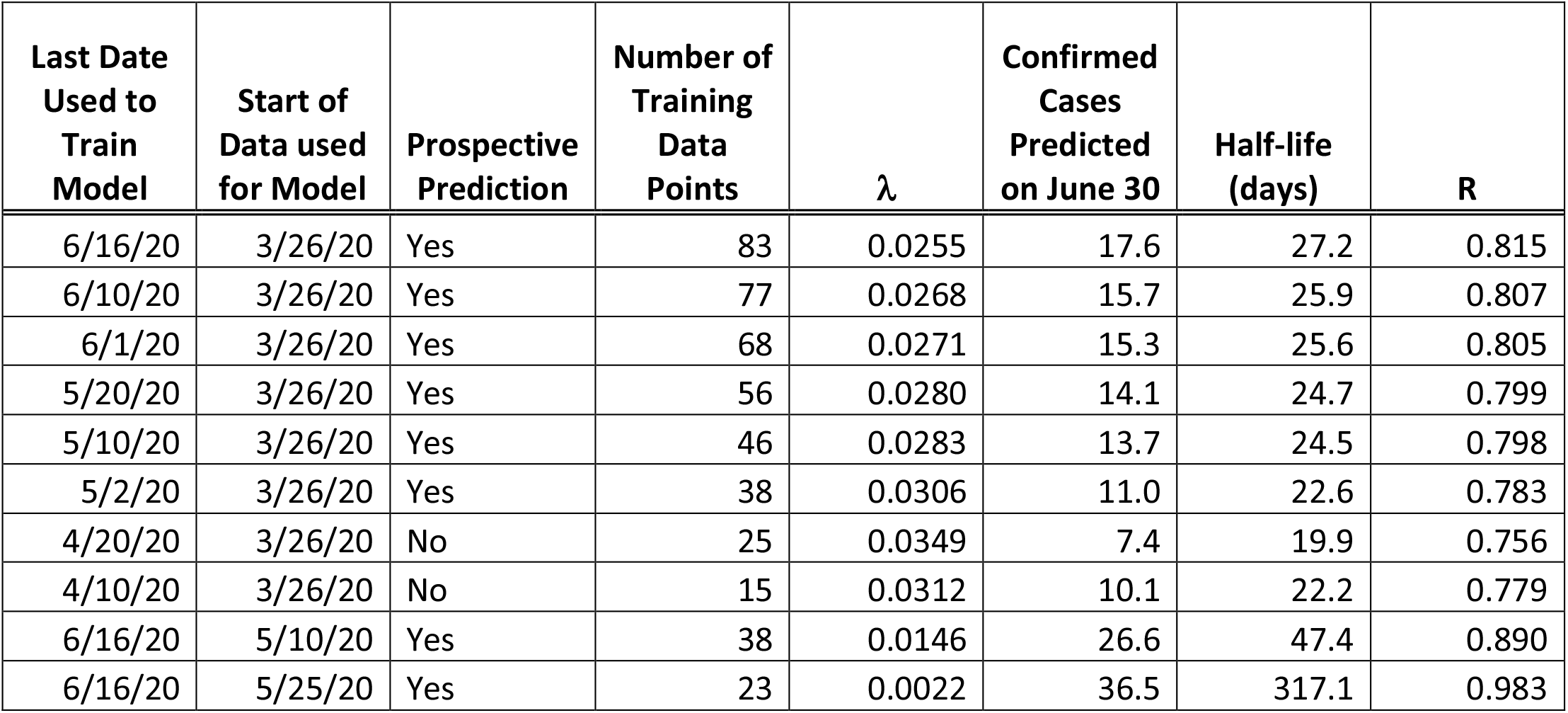
Model stability. The model’s prediction of R is relatively stable when trained over all data starting with March 26. The model was first constructed on May 2, so predictions made on dates in April are hypothetical historical predictions; they were not prospective. If the model is trained only on recent data, starting with dates in May, longer half-lives are predicted, with R approaching 1. Since data is reported by PHSKC with a one-day lag, a model with a last date of 6/16, for example, would be created on 6/17. These data suggest that R is increasing and/or that testing has enabled a higher ratio of confirmed to true case counts.

| Last Date Used to Train Model | Start of Data used for Model | Prospective Prediction | Number of Training Data Points | $\lambda$ | Confirmed Cases Predicted on June 30 | Half-life (days) | R |
| --- | --- | --- | --- | --- | --- | --- | --- |
| 6/16/20 | 3/26/20 | Yes | 83 | 0.0255 | 17.6 | 27.2 | 0.815 |
| 6/10/20 | 3/26/20 | Yes | 77 | 0.0268 | 15.7 | 25.9 | 0.807 |
| 6/1/20 | 3/26/20 | Yes | 68 | 0.0271 | 15.3 | 25.6 | 0.805 |
| 5/20/20 | 3/26/20 | Yes | 56 | 0.0280 | 14.1 | 24.7 | 0.799 |
| 5/10/20 | 3/26/20 | Yes | 46 | 0.0283 | 13.7 | 24.5 | 0.798 |
| 5/2/20 | 3/26/20 | Yes | 38 | 0.0306 | 11.0 | 22.6 | 0.783 |
| 4/20/20 | 3/26/20 | No | 25 | 0.0349 | 7.4 | 19.9 | 0.756 |
| 4/10/20 | 3/26/20 | No | 15 | 0.0312 | 10.1 | 22.2 | 0.779 |
| 6/16/20 | 5/10/20 | Yes | 38 | 0.0146 | 26.6 | 47.4 | 0.890 |
| 6/16/20 | 5/25/20 | Yes | 23 | 0.0022 | 36.5 | 317.1 | 0.983 |

### Comparison with Prior Work

By design, this exponential model is smoother than other models. This design can create some local inconsistency with other models in the context of global agreement. The model presented here has consistencies with other data and models. In particular, reports by the Seattle Flu Study (Chu et al., 2020) and the Seattle Coronavirus Assessment Network (Greater Seattle Coronavirus Assessment Network, 2020) also show data that is consistent with an exponential decay in case rate in King County over the same period.

An excellent model for the COVID-19 outbreak in King County has been developed by the Institute for Disease Modeling (IDM) (Thakkar, Burstein, Klein, and Famulare, 2020). This model is relied upon by both King County and Washington for public policy. Ideally an outbreak model should account for many hidden variables. These variables change over time. R is changing over time due to differences in population behaviors such as physical distancing. Likewise, the population is heterogeneous, and includes subpopulations such as those in dense living situations. Each of these subpopulations may contribute uniquely to a real-world model; there is no guarantee that an encompassing systems model should fit an exponential. The IDM model attempts to capture many of these variables. Furthermore, it is parameterized with the premise that these variables can change rapidly, and produce rapid and large changes in the point estimate for R. Such rapid and large changes are not typically possible to model with a two-parameter model. An additional advantage of the IDM model is that it does not require piecewise fitting. Therefore, one would expect the IDM model to better capture very short-term fluctuations in R, but to be susceptible to overfitting. Conversely, a two-parameter model limits the possibility of overfitting and should be more robust to noisy data. Consistent with this expectation, the confidence intervals for the IDM and the exponential overlap across the entire trajectory, but the point estimates for R differ considerably.

The IDM reported R=0.73 (95% CI: 0.3 to 1.2) for March 25; R=0.94 (95% CI: 0.55 to 1.33) for April 4; R=0.64 (95% CI: 0.28 to 1.0) for April 15; and R=0.89 (95% CI: 0.47 to 1.31) for April 22 (Thakkar, Burstein, Klein, Schripsema, and Famulare, 2020a; Thakkar, Burstein, Klein, Schripsema, and Famulare, 2020b; Thakkar, Burstein, Klein, and Famulare, 2020; Thakkar and Famulare, 2020). The IDM models more volatility in their point estimate for R, varying from 0.94 to 0.64 and back to 0.89 over the course of three weeks. One can replicate this volatility by adding more parameters to the empirical modeling approach, such as fitting with a local smoothing algorithm (e.g., as shown in **Figure 2**). One possible interpretation is that the IDM model has some overfitting. Since the exponential model falls within the confidence interval of the IDM model, this inconsistency may not be relevant for informing policy, as long as confidence intervals are used for reporting data, and not point estimates.

**Figure 2.**
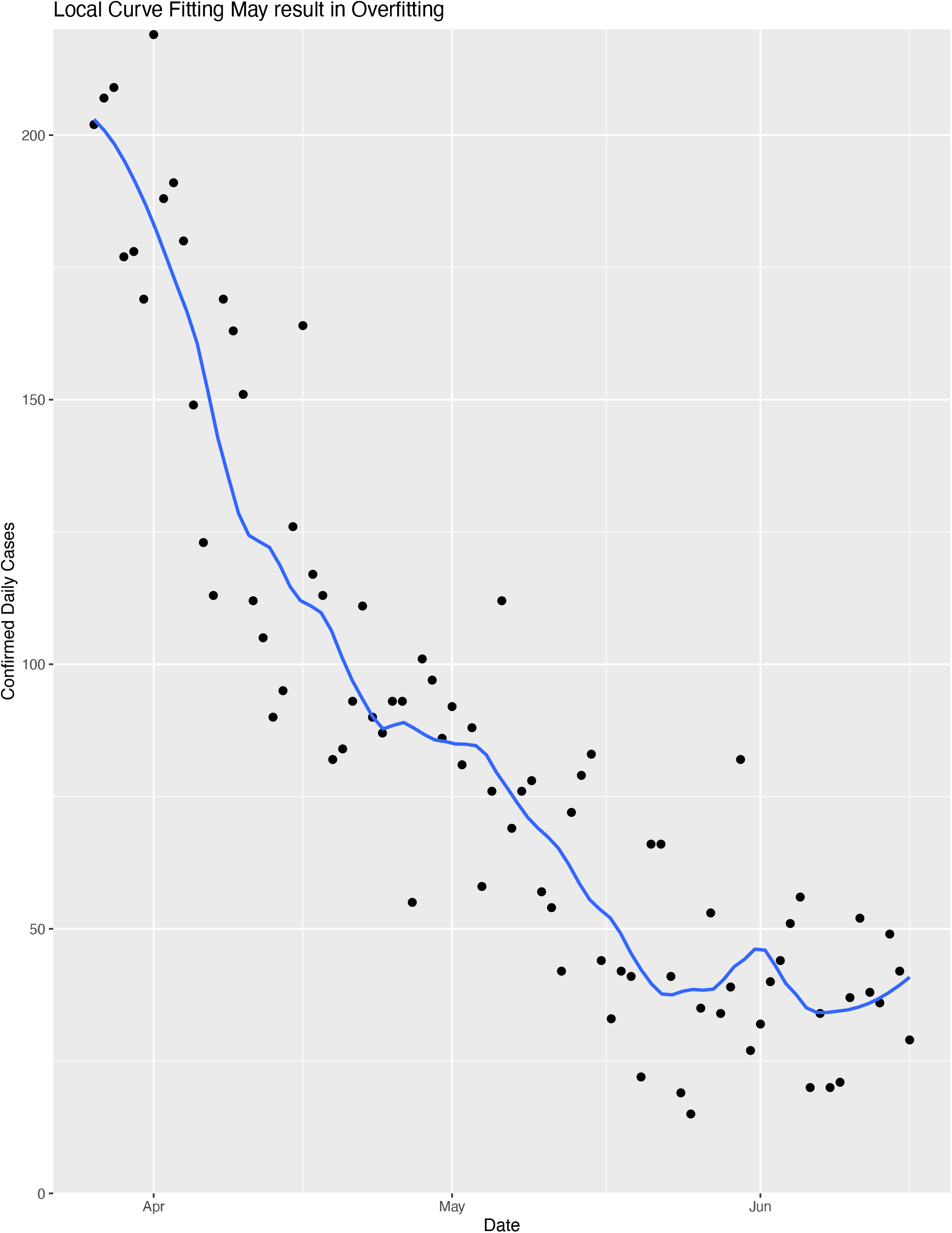
Local curve fitting may either capture daily dynamics but more likely reveals overfitting to noise. Data points are identical to those in **Figure 1**. Curve is fit with the R geom_smooth function (span=0.25), which produces a fit similar to some multiparameter models such as that of IDM. The increasing fit line at the end of June is consistent with a recently reported R=1.2 (King County, 2020a).

## DISCUSSION

Occam’s razor philosophizes that simple models are to be favored over more complex models unless there is compelling reason. Use of a simple historically accepted model limits temptation to shop for a model that fits a preconceived notion or policy. Simple models avoid overfitting to noise. Universal use of simple models increases comparability of results across studies; more complex models such as the “Susceptible, Infected and Recovered” (SIR) model are not as readily comparable as model specifications are not universal. For example, SIR models may differ in their inclusion of vital dynamics. The number of parameters can vary highly between models. For example, Wong et al. (2020) use 21 parameters; Morozova et al. (2020) use 25 parameters. To aid in comparing predictive models, the number of parameters should be prominently published with model results. In Spring 2020 in King County, there are no significant differences between the results of more complex models and a simple exponential model; the hypothesis that complex models are superior to the exponential model should be rejected.

This empirical model is sufficiently valuable that it should be used to inform public policy. It is important for public health that R be less than 1. Public policy should be crafted to keep R less than 1. If R is close to or greater than 1, more aggressive public policy measures are warranted. This model provides guidance on how close R is currently to 1. Likewise, the length of the half-life allows appropriate expectations and allocation of resources. For example, if the number of cases declines by half only every 25 days or longer, it may make sense to put less emphasis on waiting under “stay-at-home” measures for the case count to reach a much lower level and more emphasis on increasing contact tracing and other public health measures.

Use of raw reported confirmed case counts to model the real incidence of COVID19 is subject to many caveats. They undercount true case incidence, perhaps by a factor of ten. However, if this factor is relatively constant, then the estimate of half-life and R is invariant. Confirmed case counts may not uniformly sample the population. The dynamics of the outbreak(s) in King County may be substantially different in different subpopulations within the county (e.g., herd immunity may be reached in a subset of long-term care facilities). Although they have in the past produced a good fit to an exponential model (**Figure 3**), the concurrence of parameters that sums to fit an exponential model may not persist (e.g., arrow highlights in **Figure 3**). The characteristics of the population that receives tests vary over time. The number of tests performed may vary over time. If tests increase, then the true incidence may decline without a corresponding decline in confirmed incidence. The model does not currently include testing rate as a parameter. Indeed, over the March to June time frame of the model, testing rate per day has not substantially altered in King County (UW Virology COVID-19 Dashboard, 2020; PHSKC website, 2020; Washington State Department of Health website, 2020). Positive tests may be delayed by 1-2 weeks after infection. If an additional ten days embargo on the data is added, such as for the IDM model, there could be close to a month lag between a policy implementation and a statistically observable effect on the model. With extreme psychological, social, medical, and economic consequences of policy decisions, there is likely to be value in policy informed on the latest data, even if those data are subject to revision. However, policy based on just-in-time data such as the model presented here must be nimble enough to alter in the event of major model errors (like would have occurred during the week prior to 5/3/20). The data and curve fit in **Figure 1** best reflect a relatively smooth and continuous decline in cases coupled with a slowly increasing R. This is consistent with a society that is gradually decreasing effective social distancing, gradually increasing testing throughput, and gradually achieving pockets of herd immunity. These data and analyses are slightly inconsistent with an interpretation of rapid and substantial changes in R that might drive conflicting back-to-back reports from King County Public Health such as those of May 4, “COVID-19 transmission has slowed,” and May 8, “COVID-19 transmission rate could be rising in King County […] after previous indications the transmission rate had fallen below a critical threshold” (King County, 2020b).

**Figure 3.**
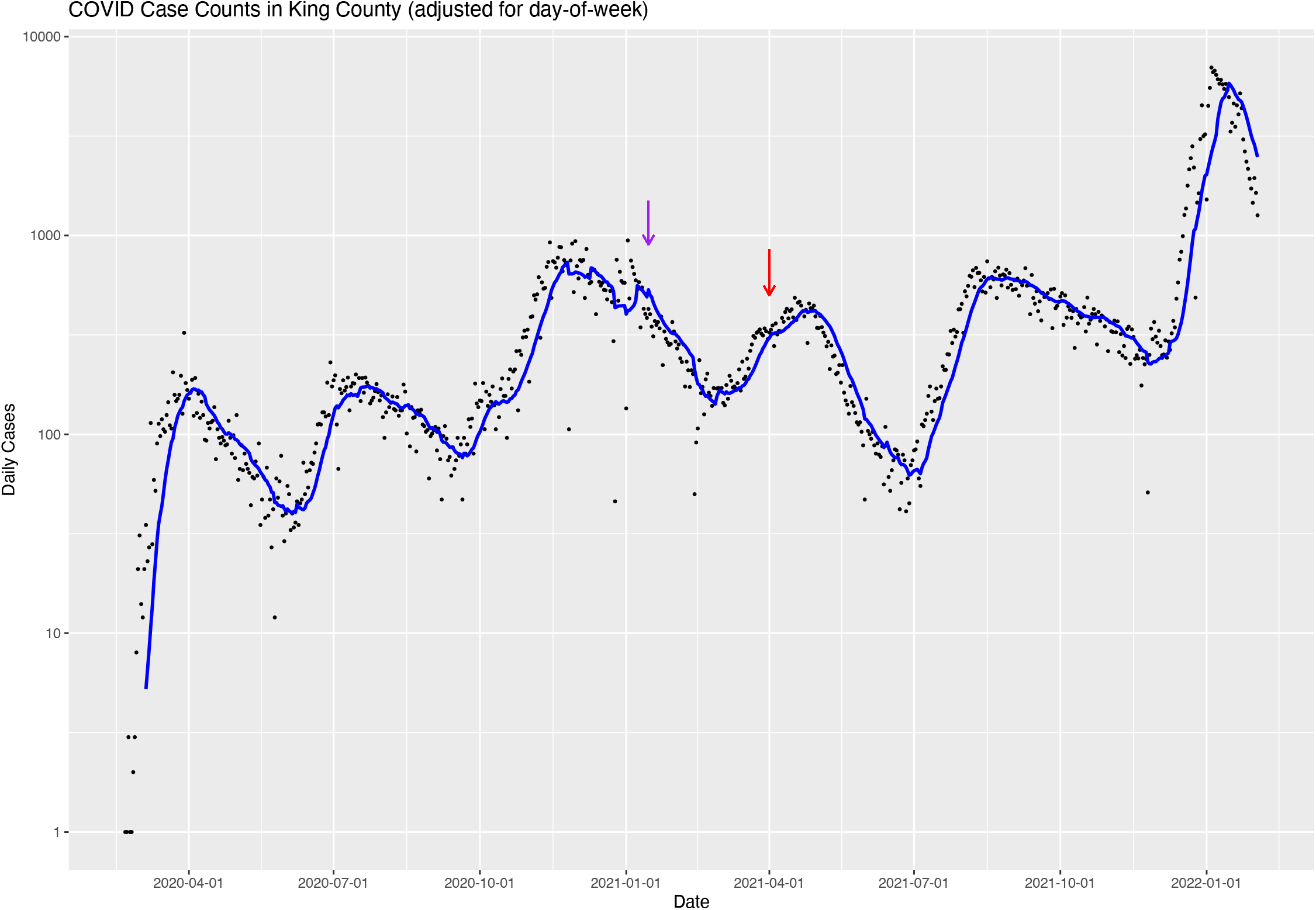
Trajectory of case counts in King County through early February of 2022. On this log-linear visualization, exponential curves appear as straight line segments. This echoes Ireland’s claim that, “If this was really the beginning of the great epidemic wave one should expect that if these series of data were plotted out on a logarithmic scale the increase from week to week would plot out as a straight line following the usual logarithmic rise of an epidemic curve” (Ireland, 1928). The entire course of the pandemic can be seen as a series of line segments, with sudden changes in slope at mostly unpredictable break points. This demonstrates that the COVID pandemic is excellently modeled as a series of exponential curves, but after varying intervals (ranging from one to four months long), the exponent of those curves will change, often dramatically. The length of these intervals is hard to predict, other than to assume future intervals will be about as long as past intervals. The majority of those changes result in switch in sign of the slope (e.g., from increasing to decreasing, or vice versa); the result is a sawtooth pattern. Two exceptions to this pattern are noted by arrows. The purple arrow points to a line segment with particularly high variability caused by snowstorms and holidays (circa New Year 2021). During these events, people shifted the dates they otherwise would have sought testing. The red arrow points to a period of high vaccination rate (circa April 2021), causing a prolonged period of increasing herd immunity, and thus substantial deviation from exponential behavior (resulting in a downward concave shape to the visualization of this interval).

Public policy should be to make as much data available to inform these just-in-time models as can reasonably be balanced against civil liberties and privacy considerations. In particular an understanding of what subpopulations are reporting case counts would considerably improve the value of these models for informing policy. It would be valuable for government agencies to produce and make available data including occupation and living environment to improve these models in a manner that appropriately protects privacy.

The “incidence count half-life (*t*1/2)” metric for post-peak outbreak modeling may be a more useful metric for communicating with the popular press than Re or R0. To date, R has largely been an academic statistic, and may be more useful for communicating approximations about the biology and mechanisms of transmission of a pathogen than for formulating public health policy. It is particularly awkward and counterintuitive that a parameter with a subscript of 0 (naught) is not a constant but is highly variable and dependent on many observed and latent parameters. R is a mathematical paralog to the statistic ‘average lifetime’ (τ) used by physicists to describe radioactive decay. In that field, τ is relatively deprecated compared to *t*1/2; *t*1/2 has many times greater usage in general and public communication than τ. Even if the relationship between R and half-life is modeled using a single parameter, the number of infectious days (*d)*,

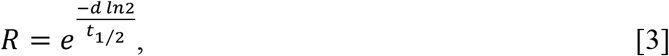

a high uncertainty in R remains unavoidable because there is considerable uncertainty in *d*. This parameter *d* may vary between individuals and may vary considerably over time for each individual. The length of time an individual is infectious and how individual infectiousness varies over time and circumstances is not known. Therefore, there is more uncertainty for estimates of R than for half-life. Half-life depends only on the observed data; therefore, a good estimate of the uncertainty in half-life can be determined from the observed case-count data. This is not true for R. Furthermore, reproduction number parameters (e.g., Re and R0) depend on many other factors that are typically incomparable between publications without expert interpretation and are neither well observed nor well known for SARS-CoV-2 (Delamater et al., 2019; Viceconte & Petrosillo, 2020). Since these factors must be estimated, a large uncertainty for R must be reported. This uncertainty in R overstates the amount of uncertainty relevant to at least some public health policy decisions. Furthermore, extra care should be used in reporting confidence intervals for R that have values near 1 or that include 1. Crossing this threshold has a large impact on public policy.

A philosophy of this approach – the use of an exponential model to describe a disease outbreak – is to keep the model as simple as possible to avoid overfitting and to avoid assuming too much knowledge about the underlying processes and parameters of the outbreak. Major changes in factors that alter the biological plausibility of the model will invalidate its predictions. These changes that have overwhelming impact on model parameters may include (1) rapid changes in herd immunity, such as occur near the peak incidence of an epidemic, (2) public policy shifts, such as “shelter in place”, that are widely adopted, (3) advent of a novel variant of concern or change, and (4) changes in human behavior. Therefore, the exponential model is best used piecewise, with separate parameter fits to intervals punctuated by such events (**Figure 3**), as recommended by Faranda et al. (2020), “Dynamical and statistical modeling should focus on limited stages of the epidemics and restrict the analysis to specific regions, thus accounting for large uncertainties.” The model will work equally well in piecewise intervals with rising or falling daily case counts. Similarly, Read et al. (2020) suggest that dynamical and statistical modeling should focus on limited stages of the epidemics and restrict the analysis to specific regions. Extrapolation based on a simple epidemiological model cannot account for rare unpredictable events. Thus, a limited focus avoids the appearance of overconfidence.

### Notes added in Proof

#### Note 1 (June 2020)

Following the data freeze analyzed in this report and the ensconcement of these predictions in MedRxiv, an inflection occurred in the trajectory of daily confirmed case counts in King County. The local minimum occurred between June 6 and June 16. This minimum occurred approximately 5-8 days (the expected lag time between changes in R and changes in confirmed case counts) after King County made a major public health policy change from “Phase 1” to “Phase 1.5” on June 5 (King County, 2020a). One concludes that this several-month period from March 26 to June 10 between major public policy changes was well modeled by a simple exponential fit, and therefore that piecewise modeling of epidemics between major policy changes with separate exponential fits for each interval is a viable – and perhaps even the best – strategy, as long as the prevalence of herd immunity during these intervals is relatively constant.

### Note 2 (February 2022)

This paper was originally published before the second wave of COVID in King County. The following waves were also well modeled with a series of exponential curves (**Figure 3**).

## Data Availability

All data was obtained from the King County Department of Health website. Data is provided as a supplemental file.

https://www.kingcounty.gov/depts/health/covid-19/data/daily-summary.aspx

## Acknowledgements

Sui Huang and Noa Rappaport provided useful comments and criticisms.

## Abbreviations

IDM: Institute for Disease Modeling
PHSKC: Public Health — Seattle & King County
R: reproduction number
R2: coefficient of determination
SIR: Susceptible, Infected and Recovered

## METHODS AND SUPPLEMENTARY NOTES

### Code

All analyses were performed with R version 4.0.1 (R Foundation for Statistical Computing, Vienna, Austria). R code is provided as a supplemental file.

### Data

Data is provided as a supplemental file. Data was acquired by manual transcription of the daily confirmed case counts daily from February 28 through June 17, 2020; more automated download was not available (PHSKC website, 2020).

One approach to best capturing the information in these variables is to use the most up-to-date, latest data. Although such data may be more incomplete and less reliable than an earlier data freeze, it may lead to a more useful model by better capturing the latest effects of dynamic changes in hidden variables. Over time, Public Health — Seattle & King County (PHSKC) improves previously reported confirmed case counts by removing duplicates, correcting residency information, adding newly received counts from previous dates, improving cause of death information, and other data cleaning. This creates a potential trade-off between using data that includes the last few reported days and using only older more reliable data. However, for a two-parameter model with many data points, inclusion or exclusion of the last two days of data has little effect on the model. Therefore, the model fit presented here includes all data, to best capture recent changes in underlying parameters, such as changes in population behavior or resolving flares in relatively isolated subpopulations, but at the risk of being more sensitive to issues associated with delayed case reporting and database maintenance.

To date, revisions to the PHSKC daily confirmed case reports fall into three categories: (1) additions to the last several days as new cases arrive in the database, (2) minor revisions to cases counts up to several weeks old, and (3) major revisions to the latest ten days of data. Revisions of the first type tend to have only a minor impact on estimates because they typically only have a noticeable magnitude of change for the last datapoint. They can also be adjusted statistically to add the expected proportion of delayed cases based on historically reported second-day adjustments. Revisions of the second type have negligible impact on estimates, as they seldom alter the counts for a given day by more than one or two counts. The third type of revision has happened only once, over the weekend, between May 2 and May 3, 2020; on May 3 a total of 105 cases counts were subtracted from the previous ten days, biased towards more recent dates, with over 20 counts being subtracted from previously reported counts for April 28, April 29, and April 30. This revision would have substantially altered the conclusions of the approach presented here and represent a major caveat to the interpretation and use of these results; these results will be substantially misleading if major anomalies occur again.

As this model intends to capture early indicators of outbreak progression, it focuses on reported cases counts, not deaths or hospitalizations. Deaths and hospitalizations, although more reliably assayed and reported and so are used in models such as that of the IHME (IHME COVID-19 Health Service Utilization Forecasting Team, 2020), are lagging indicators compared to incident case reports, but are excellent for predicting peak usage of resources.

The model presented here uses data from the PHSKC website. The IDM model uses data obtained directly from WA state. Although the PHSKC website states that it is an exact reflection of state data, there are differences between these data sets. The differences are minor and do not affect major conclusions. The reasons for these differences are not known.

### Starting Date

This model is robust to choice of initial data date. For example, for an analysis performed on May 11, choices of start date from March 26 to March 31 all result in a half-life between 24 and 25 days. Choices of later start dates produce longer half-lives, suggesting that the rate of decline may be slowing, however, uncertainty in these estimates rises as they are derived from fewer data points. For example, as of May 11, the half-life estimated from April 10 data and on was 32 days. This also suggests a slowing of the rate of decline during the month of April and early May. The choice of starting date for data fit can be considered a parameter, so it is reasonable to consider the exponential model a three-parameter model – but if that choice is made for comparative purposes then other models should also have data-selection parameters enumerated. If the exponential model were applied to historical datasets, then choice of the end date for the modeled interval would constitute a fourth parameter. Extreme changes in initial date can change model predictions (e.g., **Table 1**, bottom two rows), and suggest a change in R across different time intervals.

### Model Fit

The absolute residuals of the model are fairly constant and show little structure. This suggests that data deviating from the model fit is well described as “noise,” rather than some systematic effect, such as a change in public policy or public response to that policy (**Figure S1**). However, the residual on May 30 (index #66) was one of the four highest residuals; it occurred seven days after Memorial Day and could represent a response to social gathering on Memorial Day. However, that seems unlikely because the flanking days do not have similarly high residuals. There is also a mild weekly periodicity in the data, presumably corresponding to testing cycles (**Figure S2**). Therefore, smoothing over at least a week’s time is recommended; use of a two-parameter model achieves that goal.

### Utility of R as a Public Health Policy Tool When R Hovers Near Unity

The half-life statistic is particularly useful when used separately for pre- and post-peak modeling. R is a more versatile and general statistic that has value in informing policy throughout an outbreak. It is particularly useful if the value of R hovers near 1, as there are profound implications for policy depending upon which side of unity the statistic lies. If R is less than but near 1, half-life is near infinity and is inelegant as a reportable metric. Half-life is much more useful if R is consistently somewhat less than 1, and in these circumstances is a key statistic for planning for healthcare demand and tempo of economic and social adjustments. Similarly, if R is greater than 1, the doubling time statistic grows in value for public communication. Also, due to the nonlinearity of the models, the confidence intervals for R can be asymmetric around the point estimate, increasing the likelihood of misinterpretations across a chain of public communication.

### Rigor Metrics for Natural Language Processing

Sex as a biological variable data was not provided by Seattle & King County (PHSKC). Breakdown of daily data into subcategories including sex and ethnicity was not available in part due to privacy concerns related to small aggregate bin sizes (personal communication from PHSKC, May 28, 2020). This study did not require institutional review board review. This human-subjects exempt study did not require consent, randomization, or blinding. A power analysis was not applicable. No cell lines were used; no authentication was necessary.

### Key Resources Table

~~~
  Software and Algorithms
    Statement: All analyses were performed with R version 4.0.1 (2020-06-
06) (R Foundation for Statistical Computing, Vienna, Austria).
     Reagent or Resource: R
     Source: NA
     Identifier: (R Project for Statistical Computing, RRID:SCR_001905)
~~~

**Figure S1.**
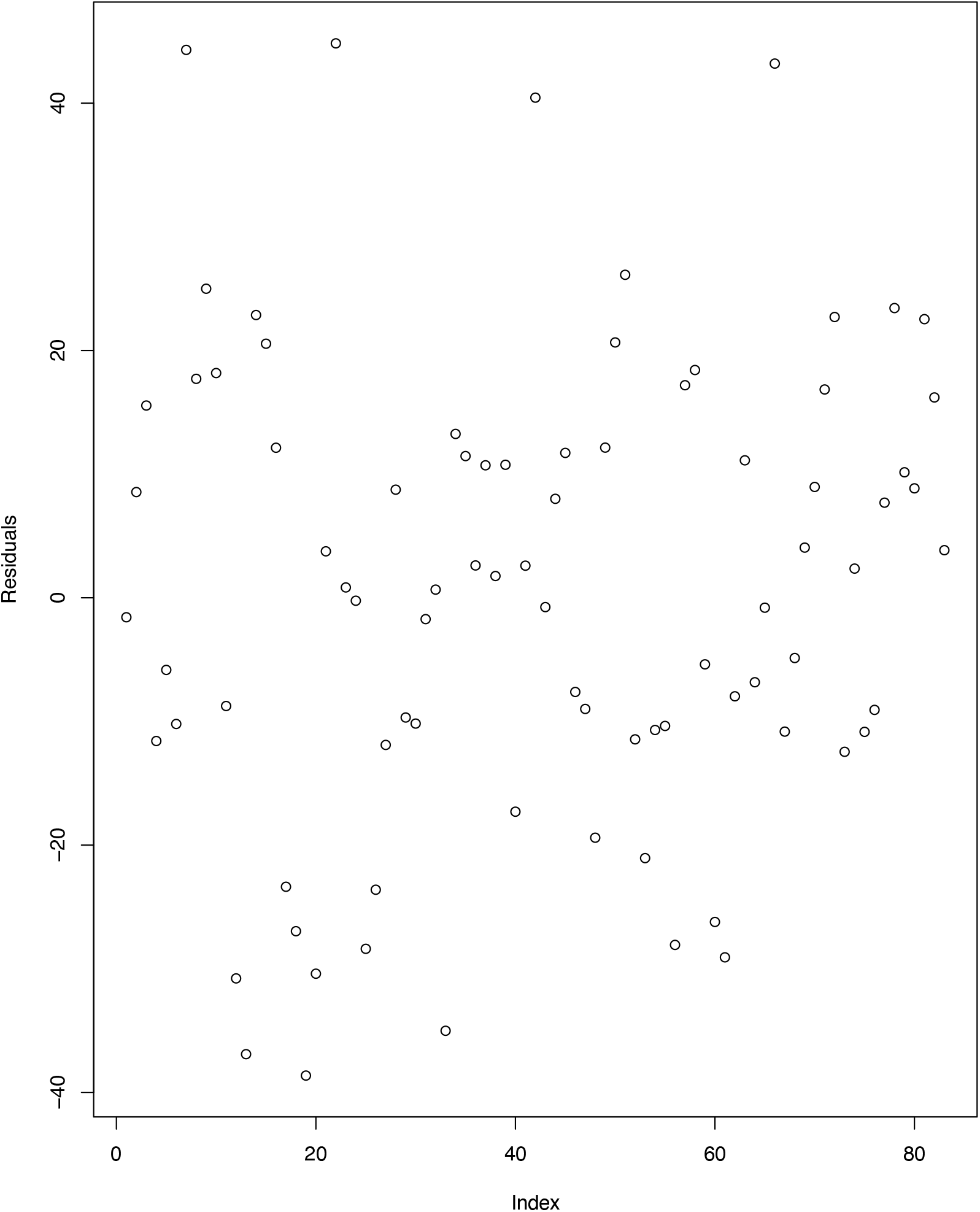
Model residuals. There is little structure to the data, and no significant structure. Therefore deviations of observed real values from model predictions are well described as noise, rather than attributed to failure of modeling assumptions.

**Figure S2.**
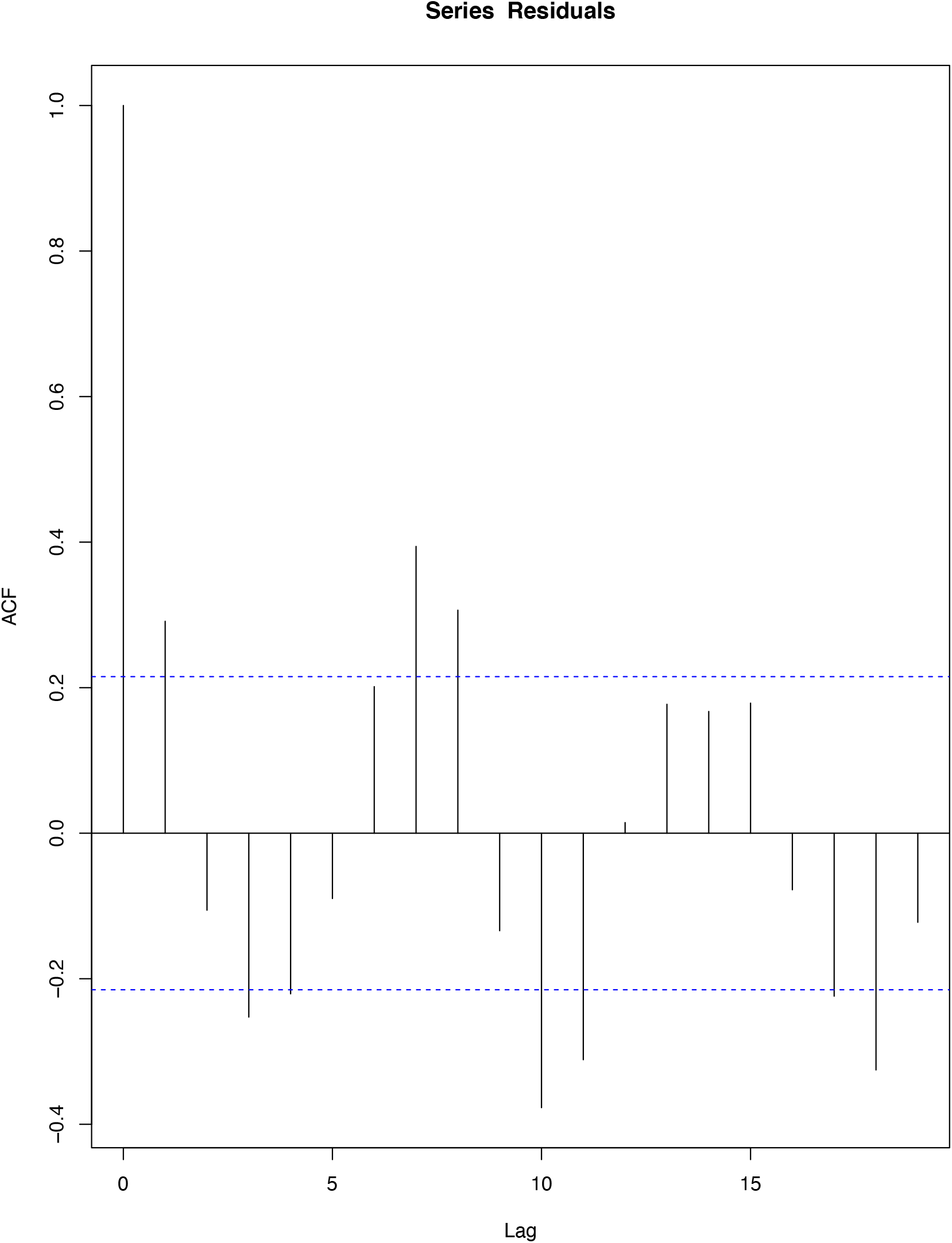
Autocorrelation of model residuals, as computed by the default parameters of the R acf function. Lag is expressed in days. A peak at seven days, and a smaller one at 14 days, indicates that there are weekly testing cycles, such as likely results from changes in availability of testing sites and in personal schedules over the weekend.

## REFERENCES

Chu HY, Englund JA, Starita LM, Famulare M, Brandstetter E, Nickerson DA, Rieder MJ, Adler A, Lacombe K, Kim AE, Graham C, Logue J, Wolf CR, Heimonen J, McCulloch DJ, Han PD, Sibley TR, Lee J, Ilcisin M, Fay K, Burstein R, Martin B, Lockwood CM, Thompson M, Lutz B, Jackson M, Hughes JP, Boeckh M, Shendure J, Bedford T; Seattle Flu Study Investigators. Early Detection of Covid-19 through a Citywide Pandemic Surveillance Platform. N Engl J Med. 2020 May 1. doi:10.1056/NEJMc2008646. PubMed PMID: 32356944; PubMed Central PMCID: PMC7206929.

Delamater PL, Street EJ, Leslie TF, Yang YT, Jacobsen KH. Complexity of the Basic Reproduction Number (R0). Emerg Infect Dis. 2019;25(1):1–4. doi:10.3201/eid2501.171901.

Faranda D, Castillo IP, Hulme O, Jezequel A, Lamb JSW, Sato Y, Thompson EL. Asymptotic estimates of SARS-CoV-2 infection counts and their sensitivity to stochastic perturbation. Chaos. 2020 May;30(5):051107. doi: 10.1063/5.0008834. PMID: 32491888; PMCID: PMC7241685.

Greater Seattle Coronavirus Assessment Network. SCAN COVID-19 Situation Report. https://publichealthinsider.com/wp-content/uploads/2020/05/SCAN_PUBLIC_SITREP-30APR-5PM.pdf. Friday May 1, 2020

IHME COVID-19 Health Service Utilization Forecasting Team, Christopher JL Murray. Forecasting the impact of the first wave of the COVID-19 pandemic on hospital demand and deaths for the USA and European Economic Area countries. medRχiv. doi: https://doi.org/10.1101/2020.04.21.20074732. 2020.

Ireland MW, editor. The Medical Department of the United States Army in the World War, vol. IX: communicable and other diseases. Washington: U.S. Government Printing Office; 1928. p. 116–7.

King County. King County Safe Start Application: Moving from Modified Phase 1 to Phase 2. 2020a. p.13

King County. Public Health news and blog. https://www.kingcounty.gov/depts/health/news.aspx. 2020b.

Morozova O, Li ZR, Crawford FW. A model for COVID-19 transmission in Connecticut. medRxiv. 2020 Jun 16. Public Health — Seattle & King County. PHSKC website. https://www.kingcounty.gov/depts/health/covid-19/data/daily-summary.aspx. 2020.

Read JM, Bridgen JR, Cummings DA, Ho A, Jewell CP. Novel coronavirus 2019-nCoV: early estimation of epidemiological parameters and epidemic predictions. MedRxiv. 2020 Jan 1.

Roach JC. Empirical Model of Spring 2020 Decrease in Daily Confirmed COVID-19 Cases in King County, Washington. MedRxiv. May 20, 2020. doi=10.1101/2020.05.11.20098798 www.medrxiv.org/content/10.1101/2020.05.11.20098798v2.

Ross. An application of the theory of probabilities to the study of a priori pathometry. Proceedings of the Royal Society A. 1915.

Thakkar and Famulare. COVID-19 transmission was likely rising through April 22 across Washington State Institute for Disease Modeling, Bellevue, Washington. May 5, 2020.

Thakkar, Burstein, Klein, and Famulare. Sustained reductions in transmission have led to declining COVID19 prevalence in King County, WA. Institute for Disease Modeling, Bellevue Washington. April 29, 2020.

Thakkar, Burstein, Klein, Schripsema, and Famulare. Physical distancing is working and still needed to prevent COVID-19 resurgence in King, Snohomish, and Pierce counties. Institute for Disease Modeling, Bellevue, Washington. April 10, 2020a.

Thakkar, Burstein, Klein, Schripsema, and Famulare. Physical distancing is working and still needed to prevent COVID-19 resurgence in King, Snohomish, and Pierce counties. Institute for Disease Modeling, Bellevue, Washington. April 10. 2020b.

UW Virology COVID-19 Dashboard. http://depts.washington.edu/labmed/covid19. 2020.

Viceconte G, Petrosillo N. COVID-19 R0: Magic number or conundrum? Infect Dis Rep. 2020 Feb 24;12(1):8516. doi: 10.4081/idr.2020.8516. PMID: 32201554; PMCID: PMC7073717.

Washington State Department of Health website. https://www.doh.wa.gov/emergencies/coronavirus. 2020.

Wong GN, Weiner ZJ, Tkachenko AV, Elbanna A, Maslov S, Goldenfeld N. Modeling COVID-19 dynamics in Illinois under nonpharmaceutical interventions. Physical Review X. 2020 Nov 16;10(4):041033.

